# Detailed Analysis of Surface Infection Barrier on Hands: Relationship with Morbidity to Infection Diseases and Identification of Antimicrobial Components

**DOI:** 10.1101/2020.12.09.20246306

**Authors:** Yuki Nishioka, Kenichi Nagano, Yoshitaka Koga, Yasuhiro Okada, Ichiro Mori, Atsuko Hayase, Takuya Mori, Kenji Manabe

## Abstract

Although the surface of hands in humans is known to harbor high levels of antimicrobial activity, reports investigating the relationship between antimicrobial activity and morbidity in infectious diseases are lacking. Additionally, the precise components involved in this activity are not known. Therefore, in this study, a method was developed to quantitatively measure the antimicrobial activity of the components found on the surface of hands, which was then compared with the medical history of the participants for infectious diseases. As a result, the antimicrobial activity of the surface of the hands was found to be positively associated with the history of infection in individuals. Furthermore, a comprehensive analysis of the components on the surface of hands indicated that organic acids and antimicrobial peptides are highly correlated with antimicrobial activity. The high amounts of lactic acid found on the surface of hands suggested it is an important factor in the hand surface infection barrier. Here we showed that the application of lactic acid within the range of the amounts present on the hand surface was found to significantly improve the antimicrobial activity of the hands. Taken together, these results demonstrate that this new method can be used to quantify antimicrobial activity, which opens new avenues for the development of hand hygiene practices by enhancing the antimicrobial activity on the surface of hands using natural ingredients against pathogens.

## 1. Introduction

Contact infection is an important route in the transmission of pathogens. Pathogens from one infected person can be transmitted either by direct contact with another person or through surfaces. Thus, contact is a major infection route for many diseases and requires control to prevent infection. Hand-washing with soap or the use of hand sanitizers with alcohol are common hand hygiene practices. In particular, the preventive effect of hand-washing on infectious diseases has been strongly demonstrated epidemiologically [1].

However, existing hand hygiene practices are unable to fully prevent contact infections, due to both the high frequency of self-inoculation and the high stability of pathogens in the environment. Regarding self-inoculation, Kwok et al. found that medical students at university touched their face an average of 23 times per hour, among which 44% came into contact with mucous membranes, such as the mouth, eyes, and nose [2]. Furthermore, in a recent survey of Iranian citizens in public places, non-mask wearers (N = 432) touched the mucous membranes of the face an average of 11 times an hour [3]. These results suggest that humans touch the mucous membranes of the face once every 5 min. Regarding the stability of pathogens, influenza virus, for example, has been reported to survive for approximately 2-8 h when normal droplets adhere to environmental surfaces [4]. In addition, the coronavirus SARS-CoV-2, which is now widespread and causing a pandemic, is even more stable than influenza and has been found to be viable for up to 72 h on environmental surfaces [5]. Therefore, it is highly possible that the virus excreted from the infected person remains on contact surfaces for a long period of time, whereby infection is caused by the frequent repetition of unknowingly touching infected surfaces and the face. In particular, in recent decades, the risk of infection by invisible pathogens has increased due to the dramatic increase in population densities and the development of transportation networks, and concerns regarding the occurrence and spread of clusters of infection, due to the unconscious and frequent transfer of pathogens from the environment to the mucous membranes of the face, are increasing.

In a meta-analysis of intervention clinical trials of hand hygiene, White et al. reported that non-alcoholic hand sanitizers using benzalkonium chloride, an antimicrobial agent, have a higher preventive effect on respiratory diseases than alcoholic hand sanitizers [6]. The viral carrier for respiratory diseases is droplets, which are more difficult to observe visually. Therefore, it is expected that unconscious self-inoculation is likely to occur in many places. The target of this survey was elementary school students, who often have a low hygiene awareness. Under these conditions, leave-on hand hygiene products with benzalkonium chloride are more likely to have a higher effectiveness for the prevention of infection compared to volatile alcohol hand sanitizers.

Hand hygiene is a common practice in daily life, wherein the excessive use of antimicrobial agents poses a risk to the development of resistant bacteria, as well taking its toll on the skin [7]. Since humans have evolved while coexisting with bacteria and viruses, it is inferred that the hands, which are closely related to infection in daily life, acquired antimicrobial ability in the process. In fact, previous studies have found that the surface of human hands has high levels of antimicrobial activity. For example, applying *E. coli* to the hands of healthy adults was found to reduce the number of bacteria by about 3 orders in 30 min [8]. Additionally, it has also been reported that the number of influenza viruses transferred to the hands decreases by 2 orders after about 5 minutes [4]. These results indicate that hands have a strong surface infection barrier, which effectively reduces bacteria and viruses, compared to other environmental surfaces. It also suggests that the identification of molecules involved in the inherent antimicrobial ability of the skin could promote the development of new natural antimicrobial agents with fewer side effects.

Several publications have proposed that the skin infection barrier involves peptides that exert antimicrobial activity and initiate the host response, resulting in the release of cytokines, inflammation, angiogenesis, and re-epithelialization [9]. Other components present on the skin, such as lipids and organic acids, may also exhibit antibacterial properties, however, their contribution to endogenous defenses remains poorly understood [10]. In addition, studies investigating the effects of the activity of the hand surface infection barrier on the morbidity of infectious diseases are lacking.

In this study, we developed a method to evaluate the inherent antimicrobial activity of the skin in combination with the quantification of organic acids, amino acids, proteins, and lipids present on the surface of hands to identify the crucial components of this barrier. Our results showed that this new method enables the precise quantification of the antimicrobial activity of the skin on the hands, which was used to successfully identify lactic acid as a crucial component of the hand surface infection barrier.

## 2. Material & Methods

### 2.1. Bacterial strains and culture medium

*Escherichia coli* NBRC3301 strain (NBRC, National Institute of Technology and Evaluation Biological Resource Center) and *Staphylococcus aureus* NBRC13276 strain (NBRC) were used. As a pre-culture, a single colony was grown on Soybean Casein Digest (SCD) agar medium (Nihon Pharmaceutical. Co., Ltd), and then inoculated into 4 mL of Luria-Bertani (LB) liquid medium (Nihon Pharmaceutical. Co., Ltd) and cultured overnight (37°C, 180 rpm). Next, 1% of the obtained culture solution was inoculated into LB liquid medium, cultured for 15 h, washed twice with sterile water, and stored on ice.

### 2.2. Measurement of viable bacterial counts by culture method using a plate reader

The viable bacterial counts were measured by monitoring growth using a Bio Microplate Reader HiTS (Sinic Corporation). A dilution series was prepared from the sample, and the bacterial numbers were measured by plating on SCD agar medium and incubating for 18 h at 37°C. Then, both the sample and the dilution series were shake-cultured at 37°C. The absorbance (OD_600_) was measured every 15 min to create a growth curve. Based on the resulting growth curve, the time required to reach OD_600_ = 0.02, which is the middle stage of the logarithmic growth phase, was obtained, and the number of viable bacteria in the sample was calculated using the calibration curve obtained from the dilution series.

### 2.3. Viral strain, cells, and culture medium

Influenza virus A/Memphis/1/71 (H3N2) was cultured and propagated in serum-free medium (SFM) (Thermo Fisher Scientific). The virus was purified by centrifugation after proliferation using Madin-Darby Canine Kidney (MDCK) cells (ATCC CCL-34). As the cells to be infected, MDCK-II cells passaged to subconfluent on a 96-well plate were used.

### 2.4. Antiviral evaluation method

A sample containing the virus was added to MDCK cells cultured on a 96-well plate. The cells were infected with the virus by incubating for 15 min at 37°C and 5% CO_2_. Then, the solution was sucked using an ejector and washed twice with phosphate buffered saline (PBS) (FUJIFILM Wako Pure Chemical Co.) at 100 µL/well. These cells were incubated at 37°C under 5% CO_2_ in SFM (100 µL/well) for 14 h, and the amount of propagated virus was calculated from the neuraminidase activity in the supernatant. Next, 20 μL of 250 µM 4-Methylumbelliferyl N-Acetyl-α-D-Neuraminic Acid (Funakoshi Co.) was added to 30 μL of supernatant and incubated at 37°C for 30 min. The reaction was terminated by the addition of 200 µL of 100 mM sodium carbonate buffer, and the fluorescence value was measured using a plate reader (Ex: 355 nm/Em: 460 nm, gain: 70) (TECAN M200). The number of viruses in the reaction solution was calculated from a calibration curve obtained from the dilution series of the virus solution measured using the same procedure. It has been confirmed that Dimethyl Sulfoxide (DMSO) has no effect on the influenza virus and MDCK-II cells under the dilution conditions used in this study.

### 2.5. *In vivo* qualitative and quantitative evaluation of surface infection barrier on hands

The antimicrobial activity was measured using a hand stamp for qualitative evaluation and a liquid cultivation method using a plate reader for quantitative evaluation. For qualitative evaluation, *E. coli s*olution (200 mL) with an optical density (OD) = 0.1 was placed in a 500 mL vat. Both hands were soaked for 5 s and the left hand was dried for 3 min. Residual bacteria were qualitatively observed by placing the hand on a hand petri dish (Nissui Pharmaceutical Co., Ltd.) containing X-Gal agar medium (Nissui Pharmaceutical Co., Ltd.) for 10 s (loading about 2 kg) and culturing in a 37°C incubator for 15 h. The other hand was stamped after drying for 30 s as a control. For quantitative evaluation, 10 µL of cultured *E. coli* solution (OD = 1.0) was applied to 4 cm^2^ on the palm of the hand, and was collected 3 min later using a swab (BD-BBL culture swab EZ; Becton Dickinson) soaked in physiological saline. Collection by swabbing was performed twice and then incubated in 1 mL of Lecithin and polysorbate 80 (LP) medium (FUJIFILM Wako Pure Chemical Co.). One hundred microliters of this sample were mixed with 100 µL of 2× SCD medium (Nihon Pharmaceutical Co., Ltd.) and cultivated in a 96-well microplate in a 37°C incubator for 24 h. Quantification was performed using a microplate reader (Bio Microplate Reader HiTS; Sinic Corporation), and the antimicrobial effect was evaluated by calculating the number of surviving bacteria relative to the initial viable bacterial number.

### 2.6. *In vitro* antimicrobial activity evaluation of hand surface components

The extraction and preparation of the hand components are shown below. A 2.5 × 5 cm region of the hand was rubbed 8 times with a swab 1K1501 (J.C.B. Industry Co.) soaked in 50% ethanol. This procedure was repeated 3 times on the same region for all subjects. Then, the swab tips were cut off, placed in a microtube, and 600 µL of 50% ethanol was added. The samples were then sonicated for 5 min using an ultrasonic cleaner SW5800 (CITIZEN Watch Co.) and pooled into one microtube (a total of 1800 µL per subject). After centrifugation at 15,000 rpm for 5 min, the supernatant was split in 2 to evaluate the antibacterial and antiviral activity. Then, the samples were dried using a centrifugal evaporator CVE-3000 (TOKYO RIKAKIKAI CO, LTD), dissolved in 10 µL of dimethyl sulfoxide (DMSO) (FUJIFILM Wako Pure Chemical Co.), and stored at −20°C. For antimicrobial activity, an undiluted solution of surface components on hands was used against *E. coli*, and an 8-fold diluted sample with PBS was used against *S. aureus*. The sample and the bacterial solution (OD_600_ = 1.0) were mixed in equal amounts and reacted at 37°C for 1 h. Quenching was performed by adding 135 µL of Lecithin and Polysorbate 80 (LP) diluent to 15 μL of the reaction solution, and then standing it on ice. Next, 100 µL of this sample was mixed with 100 µL of 2 x SCD medium and cultivated in a 96-well microplate in a 37°C incubator for 24 h, before performing quantification on a microplate reader (Bio Microplate Reader HiTS; Sinic Corporation). The antimicrobial effect was evaluated by calculating the number of surviving bacteria relative to the initial viable bacterial number. For antiviral evaluation, surface components extracted from the hand were diluted 100-fold with PBS and 100 µL were incubated with 100 µL of virus solution (8.0 × 10^4^ pfu/mL) at 37°C for 30 min. After the reaction, 100 µL of serum free medium (SFM) at a double concentration was added and placed on ice for quenching. The antiviral effect was calculated from the relative fluorescence values.

### 2.7. Blood sample collection and measurement of natural killer cell activity

Blood samples were collected in heparinized tubes with a lymphocyte preservation solution (Nipro Co.). Peripheral blood molecular cells (PBMCs) were isolated by density gradient centrifugation using Isolymph (CTL Scientific Supply Co.), according to the manufacturer’s instructions. The natural killer (NK) cell activity of the PBMCs was determined using the chromium-51 (^51^Cr) release method, as previously described [11]. Briefly, PBMCs isolated as effector cells and K-562 tumor cells labeled with ^51^Cr as target cells were incubated at a ratio of 1:50 for 4 h at 37°C. Then, the ^51^Cr released in the supernatant was measured with a γ-counter (ARC-370M, Aloka). The maximum ^51^Cr release was ensured by adding 1 N hydrochloric acid, and the minimum ^51^Cr release was confirmed by adding only complete medium to the target cells. NK cell activity was expressed as the percentage cytotoxicity, calculated using the following formula:

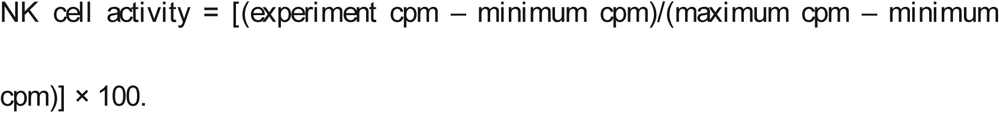

### 2.8. Saliva sample collection

Saliva samples were collected in a quiet room in the morning between 9:00 and 10:00, at least 60 min after eating. The participants were instructed to accumulate saliva in their mouths and spit it into a sterile plastic tube at the desired time. Saliva samples were collected for 10 min. The saliva volume (g) was calculated by subtracting the weight of the empty tube from the collected tube.

### 2.9. Measurement of oral mucosal moisture

The oral mucosal moisture was measured using an oral moisture-checking device (Mucus, Life Co.), performed according to the manufacturer’s instructions. Briefly, the sensor surface of the device was placed in vertical contact with the lingual mucosa, and the pressure of the sensor was controlled to approximately 200 mg. During the measurement, the participants were instructed to put their tongue out. The measurements were conducted three times, and the mean value was calculated.

### 2.10. Measurement of nasal mucociliary clearance

The nasal mucociliary clearance time was evaluated using the saccharine test, as previously described [10]. A tablet was placed in the nasal cavity of the participants, who were instructed to settle into a comfortable sitting position and not sneeze, cough, sniffle, blow their nose, talk, or take deep breaths during the test [12]. The time taken from the placement of the tablet to the perception of sweet taste was recorded as the nasal mucociliary clearance time.

### 2.11. Sample collection method for hand surface component analysis

For the collection of water-soluble components, 9 sheets of φ2.1 cm filter paper (AS ONE Co.) were attached to the palm of the left hand for 5 min, and 60 µL of ultrapure water was added dropwise to each filter paper. The filter paper was divided into 3 sheets (for low-molecular-weight compounds) and 6 sheets (for antimicrobial peptides), and each was collected in a screw tube. For lipid collection on the skin surface, a 5 × 5 cm area of the left palm was scraped with a swab soaked in ethanol (three times from the same site) and collected into a screw tube.

#### 2.11.1. Analysis of low molecular weight water soluble components on the skin surface

Three filter papers sampled from the hand surface were dried by spraying with nitrogen. Subsequently, ultrapure water was added, and the supernatant was subjected to liquid chromatography mass spectrometry. An AQUITY LC system (Waters) equipped with LTQ Orbitrap Velos Pro (Thermo Fisher Scientific) was used, and samples were separated using a Discovery HS F5 column (3 μm beads, 2.1 mm i.d., 150 mm long) (Sigma-Aldrich) at 0.2 mL/min. The elution buffer was composed of buffer A (0.1% formic acid) and buffer B (acetonitrile containing 0.1% formic acid). The mobile phases were programmed consecutively, as follows: an isocratic elution of A 95% (B 5%) between 0 and 5 min, a linear gradient of A 95-0% (B 5-100%) between 5 and 20 min, an isocratic elution of A 0% (B 100%) between 20 and 25, and an isocratic elution of A 95% (B 5%) from 25.1 to 40 min for column equilibrium. The scan measurements were performed in positive and negative ion mode.

#### 2.11.2. Analysis of water-soluble proteins on the skin surface

The identification of superficial water-soluble proteins was carried out as previously described [13]. Briefly, protein extracts from 6 filter papers were dissolved in MPEX PTS Reagents (GL Sciences) containing 7 M urea and 2 M thiourea. Then, 1/100 amount of 50 mM ammonium bicarbonate (NH_4_HCO_3_) buffer containing 1 M dithiothreitol was added to the sample solution, and the mixture was incubated for 16 h at 35°C. Subsequently, 1/20 amount of 50 mM NH_4_HCO_3_ buffer containing 1 M iodoacetamide was added to the sample solution, and the mixture was incubated for 30 min at room temperature. The amount of protein was quantified using the EZQ protein assay kit (Thermo Fisher Scientific). The solution was diluted 3 times with 50 mM NH_4_HCO_3_ before degrading using lysyl endopeptidase (FUJIFILM Wako Pure Chemical Co.) and trypsin (FUJIFILM Wako Pure Chemical Co.). The amount of lysyl endopeptidase and trypsin was 1/100 of the amount of protein in each sample. Then, ethyl acetate and trifluoroacetic acid (0.5% of final concentration) were added to the mixture. Then, after centrifugation at 15,700 × *g* for 2 min, the supernatant was removed and dried under reduced pressure at 50°C to obtain a pellet. The resulting pellet was dissolved in water containing 0.1% trifluoroacetic acid and 5% acetonitrile and applied to the GL-Tip SDB column (GL Sciences, Tokyo). After washing once with water containing 0.1% trifluoroacetic acid and 5% acetonitrile, it was eluted with 80% acetonitrile containing 0.1% trifluoroacetic acid. The eluate was dried under reduced pressure at 35°C, dissolved in water containing 0.1% formic acid and 2% acetonitrile, and subjected to liquid chromatography tandem-mass spectrometry. The samples were concentrated with Acclaim PepMap 100 C18 Nano-TrapColumn (3 μm beads, 75 μm id, 20 mm long; Thermo Fisher Scientific) and separated using a nanoACQUITY BEH130 C18 (1.7 μm beads, 100 μm id, 100 mm long; Waters). Proteins were identified by cross-referencing the obtained spectrum with the Swiss-Prot protein sequence database. The concentration of the identified protein was estimated using the emPAI method [14].

#### 2.11.3. Analysis of lipids on the skin surface

Superficial lipids were quantified as previously described [15]. After immersing a cotton swab with scraped hands in chloroform/methanol (1/1 solution), the supernatant was collected and internal standard substances (TAG-C39: 0: tritridecanoin, DAG-C26: 0: ditridecanoin, FFA-C12: 0-d3: lauric acid (Methyl-d3), WE-C28: 1: lauryl palmitoleate, ChE-C10: 0: cholesteryl caprate, ChE-C2: 0:cholesteryl acetate) were added to samples that were then subjected to liquid chromatography tandem-mass spectrometry. An Agilent 1200 series LC system equipped with an Agilent 6460 set ESI source was used. Chloroform/methanol (50:50, v/v) containing 15 mmol/L ammonium acetate was used as the mobile phase and analyzed at a flow rate of 0.2 mL. Quantification was performed using the internal standard method. For the quantification of compounds of different types and chain lengths, differences in intermolecular sensitivity were as corrected from the relative molar sensitivity with the internal standard substance, and the absolute quantification value was calculated.

## 2.12. Study design

The following four surveys were conducted regarding the surface infection barrier of the hand: (1) a preliminary study to understand the antimicrobial activity of hands using *in vivo* and *in vitro* methods; (2) a retrospective study investigating the relationship between antimicrobial activity of hand and morbidity to infection; (3) the identification of antimicrobial components on hands using comprehensive analysis; (4) the verification of the effect of lactic acid application.

### 2.12.1. A preliminary study to understand the antimicrobial activity of hands using *in vivo* and *in vitro* methods

This study was conducted as a pilot study, and volunteers were recruited randomly. The subjects were healthy adult men and women (aged 25 to 49 years). The participants performed standard hand washing with test soap formulated with alkyl ether carboxylic acid, sodium salt (AEC), alkyl ether sulfate (AES), and alkyl glucoside (AG) without antimicrobial compounds [16], rinsed with tap water for 30 seconds, and then washed using purified water for 10 seconds. To avoid contact with the evaluation site, they wore polyethylene gloves (AS ONE Co.) for 2 h of acclimation. Subsequently, the qualitative and quantitative evaluation of the antimicrobial ability of the hand was performed according to the method described separately.

### 2.12.2. A retrospective study investigating the relationship between the antimicrobial activity of hands and morbidity to infection

The relationship between the antimicrobial activity of hands, other physiological functions known to act against infectious diseases (saliva volume, oral mucosal moisture, NK cell activity in PBMCs, and nasal mucociliary clearance), and morbidity to infection was investigated in a retrospective study. This survey was conducted in June 2018. Thirty-eight healthy men and 71 women between the ages of 20 and 49 years who lived with children under elementary school age were recruited. Based on the responses to questionnaires on infectious diseases (Table S1), we constructed 2 groups: (1) a “high morbidity” group, comprised of 55 subjects; (2) a “low morbidity” group, comprised of 54 subjects (Table S1). The restrictions on the subjects were as follows: (1) avoid excessive drinking, eating, and strenuous exercise the day before the test; (2) smoking is prohibited on the day of the test until the end of the test; (3) have breakfast and brush teeth for at least 1 hour before starting the test; (4) only drink water from 1 hour before starting the test; (5) avoiding washing hands during the test.

In addition to measuring the biological parameters, a lifestyle-related questionnaire was also provided (Table S2). Statistical analysis was performed using the Statistical Package for the Social Sciences (SPSS) software for Windows. The data were first tested for normality using the Shapiro–Wilk test. Data showing normality were assessed for variability using an f-test and t-test, in accordance with variability (body temperature, NK cell activity in PBMCs, and nasal mucociliary clearance). Data that did not show normality were tested using the Mann-Whitney U test (saliva volume, oral mucosa moisture, and antimicrobial activity of hand for *E. coli* and *S. aureus*). In addition, we performed decision tree analysis to assess the biological parameters (age, sex, and body temperature) and physiological functions (antimicrobial activity of hand, saliva volume, oral mucosal moisture, NK cell activity in PBMCs, and nasal mucociliary clearance) associated with morbidity to infection. In this analysis, we start with a core node consisting of the entire sample, and then recursively partition each node into two child nodes to create a tree-like structure. Odds ratios (ORs) and 95% confidence intervals (CIs) were obtained using the cross-tabulation method.

### 2.12.3. Identification of antimicrobial components on hands using comprehensive analysis

The participants were 27 healthy men and 27 women aged 20 to 49 years, resulting in a total of 54 subjects. The test was conducted in January 2018. Subjects performed standard hand washing with test soap formulated with AEC, AES, and AG without antimicrobial compounds [16], rinsed with tap water for 30 seconds, and then washed using purified water for 10 seconds. To avoid contact with the evaluation site, they wore polyethylene gloves (0950; SHOWA GLOVE Co.) for 2 h of acclimation (20°C, 40% humidity). The hand components were collected at the sites shown in Figure S1. The correlation coefficient between each compound and antimicrobial activity was calculated using the corr.test function in R.

### 2.12.4. Verification of the effect of lactic acid application on the antimicrobial activity of hands

A test site of 2 × 2 cm on the palm of the subjects’ hands was used. After hand washing, as described above, 10 µL of 0.2, 1.0, 5.0, 10, and 23.5 g/L lactic acid aqueous solution (FUJIFILM Wako Pure Chemical Corporation) was added dropwise to the test site, spread over 30 s using a 0.2 mL round tube (AS ONE Co.), and dried for 5 min. During this process, 5.6, 27.8, 138.8, 277.5, and 652.5 nmoL/cm^2^ of lactic acid remained on the hands, respectively. Next, 10 µL of the bacterial solution (*E. coli*, OD = 1) was added dropwise to the test site, spread for 30 s using a 0.2 mL round tube, and dried for 3 min. The test site was scraped with a swab immersed in LP-PBS containing PBS with LP solution to collect the residual bacteria. Collection by swabbing was performed twice and suspended in 1 mL of LP-PBS solution. Then, 100 µL of this sample was mixed with 100 µL of 2× SCD medium (Nihon Pharmaceutical Co.) and cultivated in a 96-well microplate in a 37°C incubator for 24 hours, followed by quantification on a microplate reader (Bio Microplate Reader HiTS; Sinic Corporation). To determine if the effect of lactic acid is mediated by pH, we used aqueous hydrochloric acid (HCl) solution (Kanto Chemical Co., Inc.), in which the pH was adjusted to the same value of lactic acid as the control.

## 3. Results

### 3.1. Preliminary study to characterize the antimicrobial activity of hands using *in vivo* and *in vitro* methods

As a first pilot study, we examined various methods for measuring the antimicrobial activity found on hands. After immersing the hands of our subjects in an *E. coli* suspension, the palms were allowed to air-dry for 30 s or 3 min before being pressed against agar medium, which was cultured overnight (Figure 1A). Blue colonies indicated where *E. coli* was degrading X-gal in the medium. No blue colonies were observed in the imprints of hands not immersed in *E. coli* solution, suggesting that the microorganisms that formed the blue colonies were not found in the normal skin microbiota (data not shown). As shown in Figure 1A, regarding subject (a), the number of bacteria decreased remarkably in 3 min compared to the control (30 s), indicating the ability of the human hand to inactivate *E. coli* over time. On the other hand, regarding subject (b), no significant difference was observed between 30 s (control) and 3 min, indicating that there are individual differences in the antimicrobial ability of hands. In addition, the phenomenon of *E. coli* inactivation observed in subject (a) was not observed immediately after washing hands, suggesting that the components existing on the surface of the hands are a contributing factor (Figure S2).

**Figure 1.**
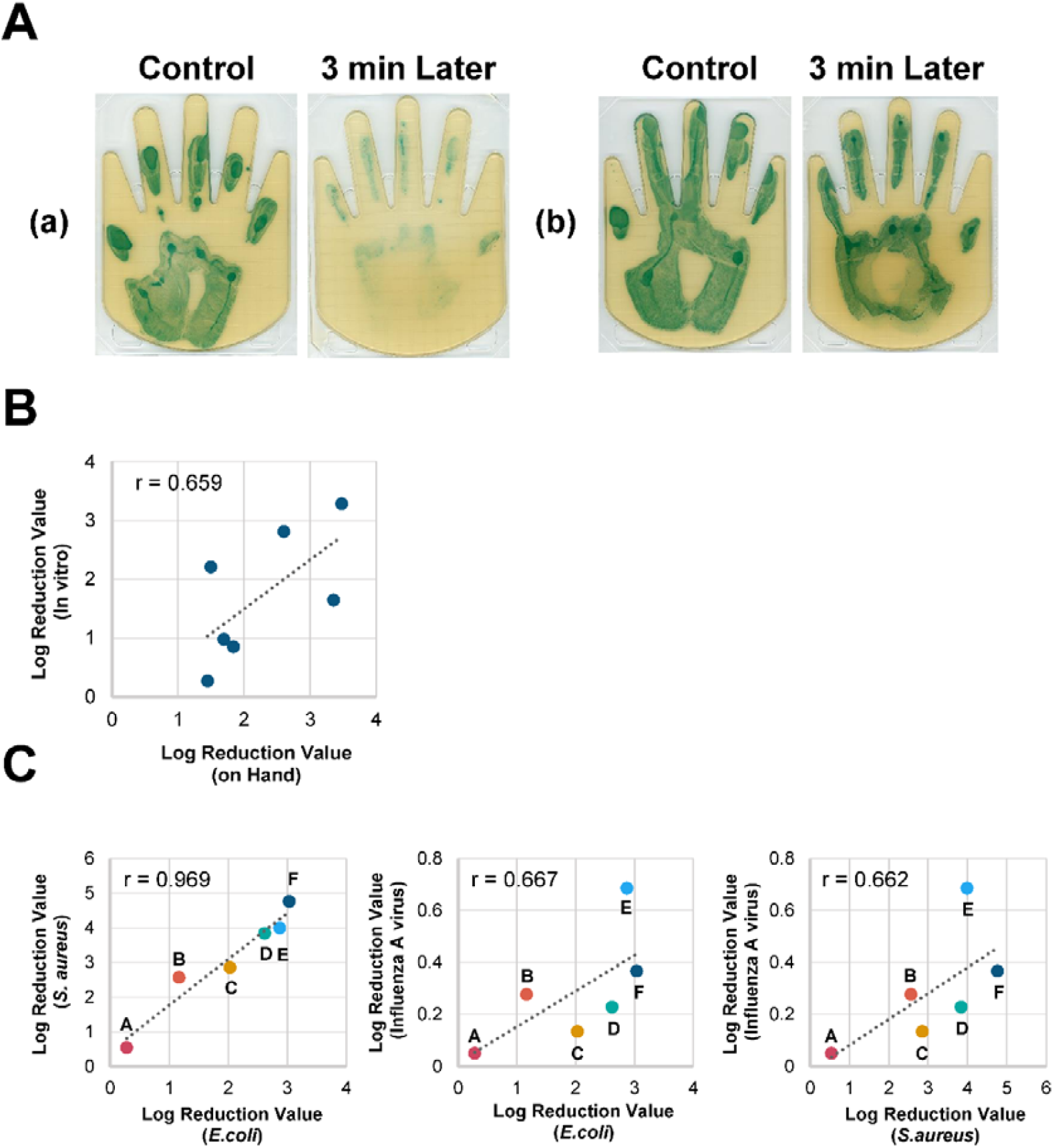
Individual differences in hand antimicrobial activity and the importance of surface components. A: Qualitative comparison results using agar medium for (a) hands with high antimicrobial activity and (b) hands with low antimicrobial activity. Results for 30 s (control) and 3 min after applying *E. coli* (OD = 0.2) to the hand. **B: Correlation between the antimicrobial activity of the hands (*in vivo*) and the antimicrobial activity of the surface components of the hands (*in vitro*)**. The antimicrobial activity of the hands was measured by applying an *E. coli* solution (OD = 0.2) and swabbing after 3 min. The surface components of the hands were collected with a cotton swab, extracted with ethanol, mixed with an equal amount of *E. coli*, and concentrated to 0.8 μL/cm^2^. The antimicrobial activity was measured by mixing the sample and an equal amount of *E. coli* (OD = 0.02) and reacting for 30 min. The log reduction value denotes the relative logarithmic reduction of viable bacteria. The value of Pearson’s correlation coefficient is indicated in the graph. **C: Correlation of the antimicrobial activity of the surface components of the hands against *E. coli, S. aureus*, and influenza A virus**. Symbols with the same color indicate individual participants between the three graphs. *E. coli, S. aureus*, and influenza A virus were evaluated using undiluted samples, 8-fold diluted samples, and 100-fold diluted samples, respectively, due to the toxicity of DMSO. The log reduction value denotes the relative logarithmic reduction of the bacterial or viral number. The value of Pearson’s correlation coefficient is indicated in each graph.

To further evaluate the antimicrobial activity of compounds on the surface of the hands, we developed a quantitative method by collecting these components and testing their effects on bacterial or viral proliferation *in vitro*. The samples were collected using a swab and extracted with 50% ethanol and dried before being concentrated to 0.8 μL/cm^2^ with DMSO. Figure 1B shows the correlation of the log reduction rate against *E. coli in vivo* method and the in *vitro* method for 7 participants, indicating a positive correlation (r = 0.66 and *P* = 0.1; Pearson correlation test). This result suggests that the components found on the surface of hands are important factors in defining the hand surface infection barrier. Furthermore, these findings indicate that this in *vitro* method is appropriate to evaluate the function of this infection barrier. Figure 1C shows the antimicrobial activity of the hand surface components of 6 participants against *E. coli* and *S. aureus*, as well as the antiviral activity against influenza A virus. For this, undiluted, 8-fold diluted, and 100-fold diluted solutions of extracted samples against *E. coli, S. aureus*, and influenza A virus were used. As a result, positive correlations (r = 0.970 and *P* = 0.001 (*E. coli* vs. *S. aureus*), r = 0.667 and *P* = 0.147 (*E. coli* vs. influenza A virus), and r = 0.662 and *P* = 0.152 (*S. aureus* vs. influenza A virus)) were observed, using the Pearson correlation test. These results suggest that the components found on the surface of hands could act on a wide range of microbial species. However, taking into account the differences in the dilution rate, the susceptibility of the hand surface components was around one order lower against *E. coli* than against *S. aureus* and influenza virus.

### 3.2. Relationship between antimicrobial activity on the surface of hands and morbidity to infection

Next, in order to assess the influence of the antimicrobial activity of hands on morbidity to infection, subjects with similar lifestyles (Table S2), corresponding to the low and high morbidity group (Table S1), were recruited and various physiological functions known to act against infectious diseases (mentioned in the Materials and Methods section) were measured, including the antimicrobial activity of the surface of hands. Figure 2 shows the antimicrobial activity of the hand surface components of each group against *E. coli* (Fig. 2A) and *S. aureus* (Fig. 2B). The antimicrobial activity against *E. coli* was significantly lower in the high morbidity group than in the low morbidity group (median of low morbidity group: 0.466; median of high morbidity group: 0.159; *P* = 0.001). Similar results were observed in the antimicrobial activity against *S. aureus* (median of low morbidity group: 1.137; median of high morbidity group: 0.191; *P* < 0.001). In addition, our study showed that not only the antimicrobial activity on hands, but also oral mucosal moisture, was associated with infection prevalence (*P* = 0.006) (Table 1). On the other hand, there was no difference between the groups in terms of their NK cell activity and saliva volume (Table 1). These results are not related to hygiene behaviors, such as hand hygiene, mask, and vaccination frequency, since the frequency of these behaviors was higher in the high morbidity group than in the low morbidity group (Table S2).

**Table 1.** Physiological functions in a retrospective study investigating the relationship between the antimicrobial activity of hands and morbidity to infection. The data were first tested for normality using the Shapiro-Wilk test. Data showing normality were assessed for variability by f-test and t-test, in accordance with variability. Data that did not show normality were tested with the Mann-Whitney U test.

|  | Low morbidity group |  |  | High morbidity group |  |  | *p*-value<sup>2</sup> |
| --- | --- | --- | --- | --- | --- | --- | --- |
|  | n<sup>1</sup> | Mean | SD | n<sup>1</sup> | Mean | SD |  |
| body temperature (°C) | 54 | 37.0 | 0.3 | 55 | 37.0 | 0.3 | 0.666 |
| saliva volume (g) | 54 | 2.4 | 1.5 | 55 | 2 | 1.4 | 0.153 |
| oral mucosal moisture (%) | 54 | 30 | 3.3 | 55 | 27.8 | 4.4 | 0.006 |
| NK cells activity in PBMCs (%) | 54 | 49.2 | 18.4 | 55 | 50.7 | 18 | 0.756 |
| nasal mucociliary clearance (sec.) | 48 | 700 | 339 | 42 | 638 | 307 | 0.371 |
| Antimicrobial activity of hand (log reduction) |  |  |  |  |  |  |  |
| for *E. coli* | 54 | 0.7 | 0.8 | 55 | 0.5 | 0.7 | 0.001 |
| for *S.aureus* | 54 | 1.5 | 1.3 | 55 | 0.9 | 1.5 | < 0.001 |

<sup>1</sup>Parameters for which the number of samples is lower than all subjects indicate the parameters for which the corresponding number of samples was not determined.
<sup>2</sup>P-values were calculated using Mann-Whitney U test.

**Figure 2.**
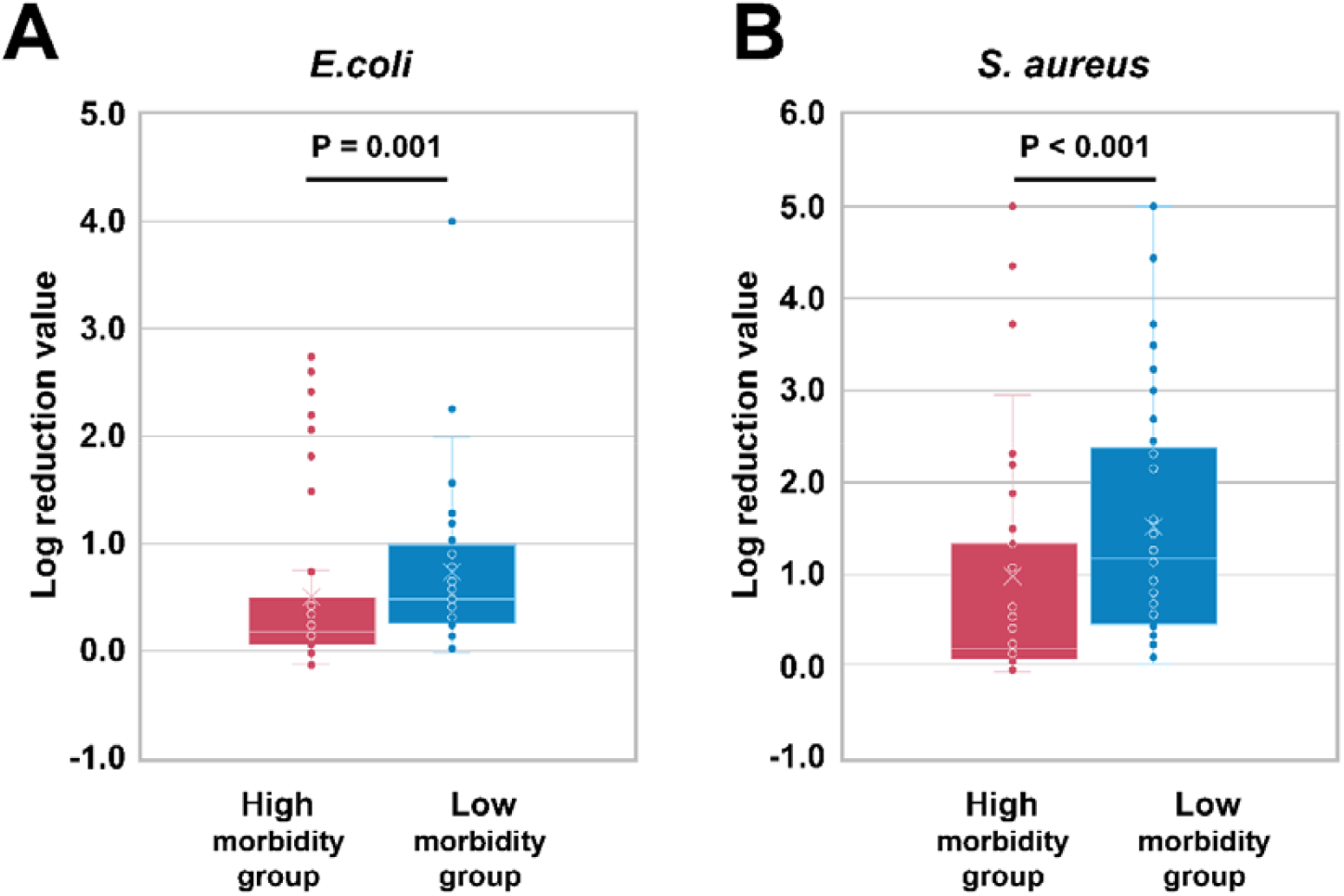
Relationship between antimicrobial activity and morbidity to infection. Result of antimicrobial activity against *E. coli* and *S. aureus*. Left (red) and right (blue) denotes the high morbidity group (N=55) and the low morbidity group (N=54), respectively. Antimicrobial activity was measured using an *in vitro* method, where the log reduction value denotes the relative logarithmic reduction of viable bacteria. Comparison between the groups was performed using the Mann-Whitney U test.

To evaluate the importance of parameters associated with infectious disease morbidity, we performed a hierarchical decision tree analysis, containing all the measured biological parameters (Figure 3). The tree comprised a total of eight nodes and five terminal nodes (nodes that have no child nodes). The antimicrobial activity of the hand (*E. coli*) was identified as the main contributing factor for morbidity to infection. The odds ratio based on the reference value (log reduction value for *E. coli*: 0.154) calculated by the decision tree analysis was 7.2 times (95% CI, 2.64-19.50). For subjects with a log reduction value for *E. coli* of 0.154 or higher, the OR based on the reference value of mucosal moisture was 5.7 times (95% CI, 2.08-15.60). With regards to age, cross-tabulation based on 41 and 38 years of age showed that the results were significant with respect to 41 years of age. However, the 95% CI ranged from 0.01-1.08, indicating that the reliability of the results was low. These results are shown in detail in Table S3.

**Figure 3.**
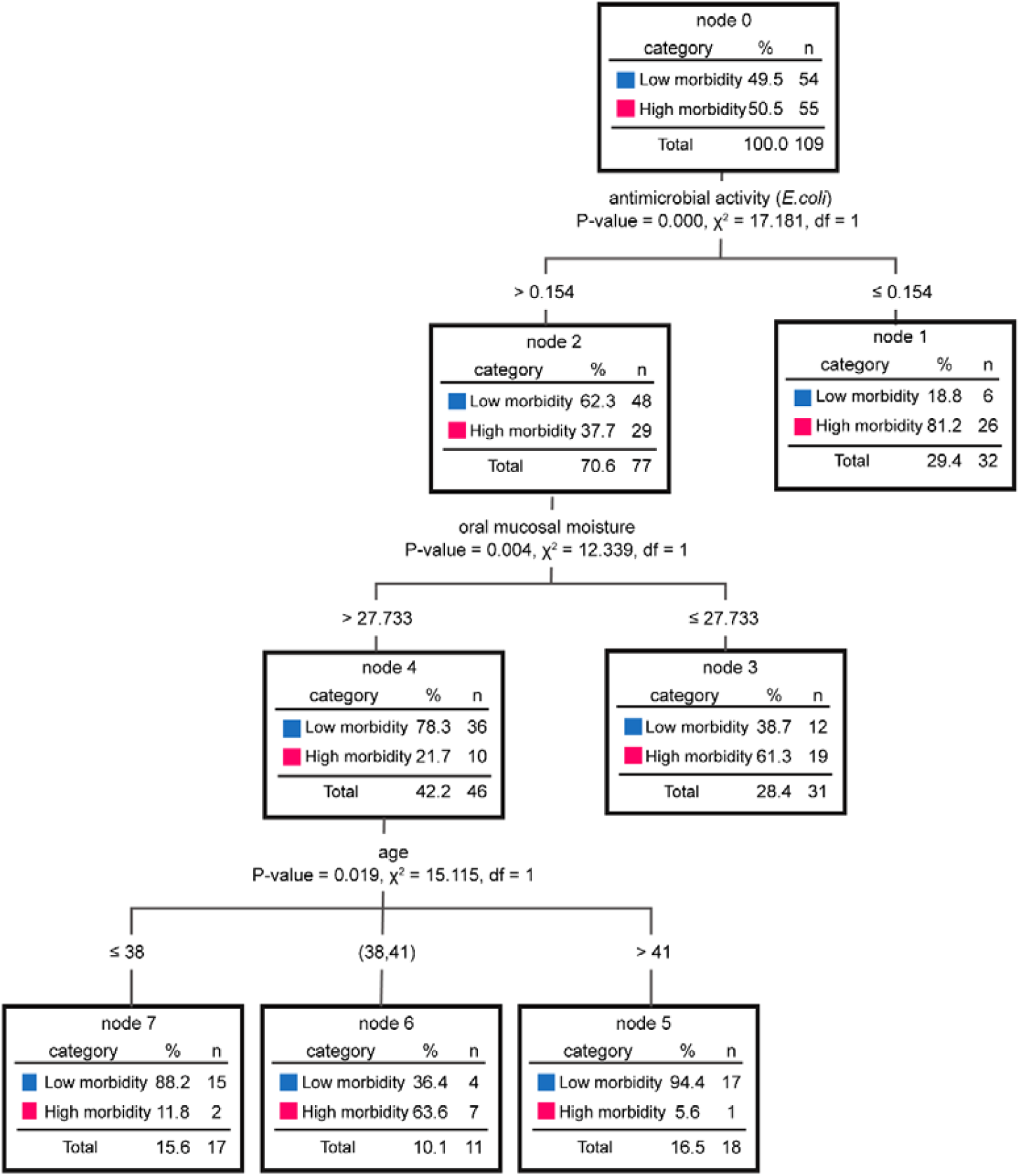
Decision tree analysis to assess the biological parameters and physiological functions. The biological parameters (age, sex, and body temperature) and physiological functions (antimicrobial activity of hand, saliva volume, oral mucosal moisture, NK cells activity in PBMCs, and nasal mucociliary clearance) were used. Odds ratio (ORs) and a 95% confidence interval were obtained from the cross-tabulation method.

### 3.3. Identification of antimicrobial components on hands using comprehensive analysis

Additionally, we conducted another clinical study to identify the factors involved in the antimicrobial activity of hands. In this survey, we analyzed the components of the surface of the hands and calculated their correlation with antimicrobial activity against *E. coli, S. aureus*, and influenza A virus. The components analyzed were organic acids, amino acids, lipids, and some antimicrobial proteins (Table 2). First, in terms of the amounts found on the hand surface, lactic acid was prominently present among all compounds (Table 2). Next, we analyzed the correlation between antimicrobial activity and surface components using Holm-corrected Pearson correlation coefficients, as shown in Table 2. Bold numbers denote statistically significant data (*P* < 0.05), while compounds in bold indicate compounds that were significantly correlated with all antimicrobial activities. Regarding organic acids, lactic acid was found to have a significant correlation with all microbial species (Table 2, Figure S3). In addition to lactic acid, malic acid, 2-hydroxybutyric acid, citric acid, ascorbic acid, azelaic acid, 2-oxoglutarate, subericacid, and fumaric acid showed a high positive correlation with antimicrobial activities. In the amino acid group, alanine, creatinine, taurine, choline, creatine, and p-aminobenzoic acid were confirmed as components with high levels of correlation. Subsequently, free fatty acids (FFA), wax ester (WE), cholesteryl ester (ChE), squalene (SQ), squalene epoxide (squalene epoxide), squalene monohydroperoxidperipheral (SQOOH), DAG, and TAG were analyzed. As a result of the measurement, small amounts of lipids were found on the hands. Among them, fatty acids were relatively abundant, among which C16: 0, C14: 0, and C12: 0 were abundantly detected (Table 2). However, none had a significant positive correlation with the antimicrobial activity of hand surface components, and only a negative correlation with DAG. Lastly, antimicrobial proteins were analyzed using shotgun proteomics analysis, including dermcidin, prolactin-inducible protein (PIP), and lysozyme C, among which prolactin-inducible protein was significantly correlated with antimicrobial activity.

**Table 2.**
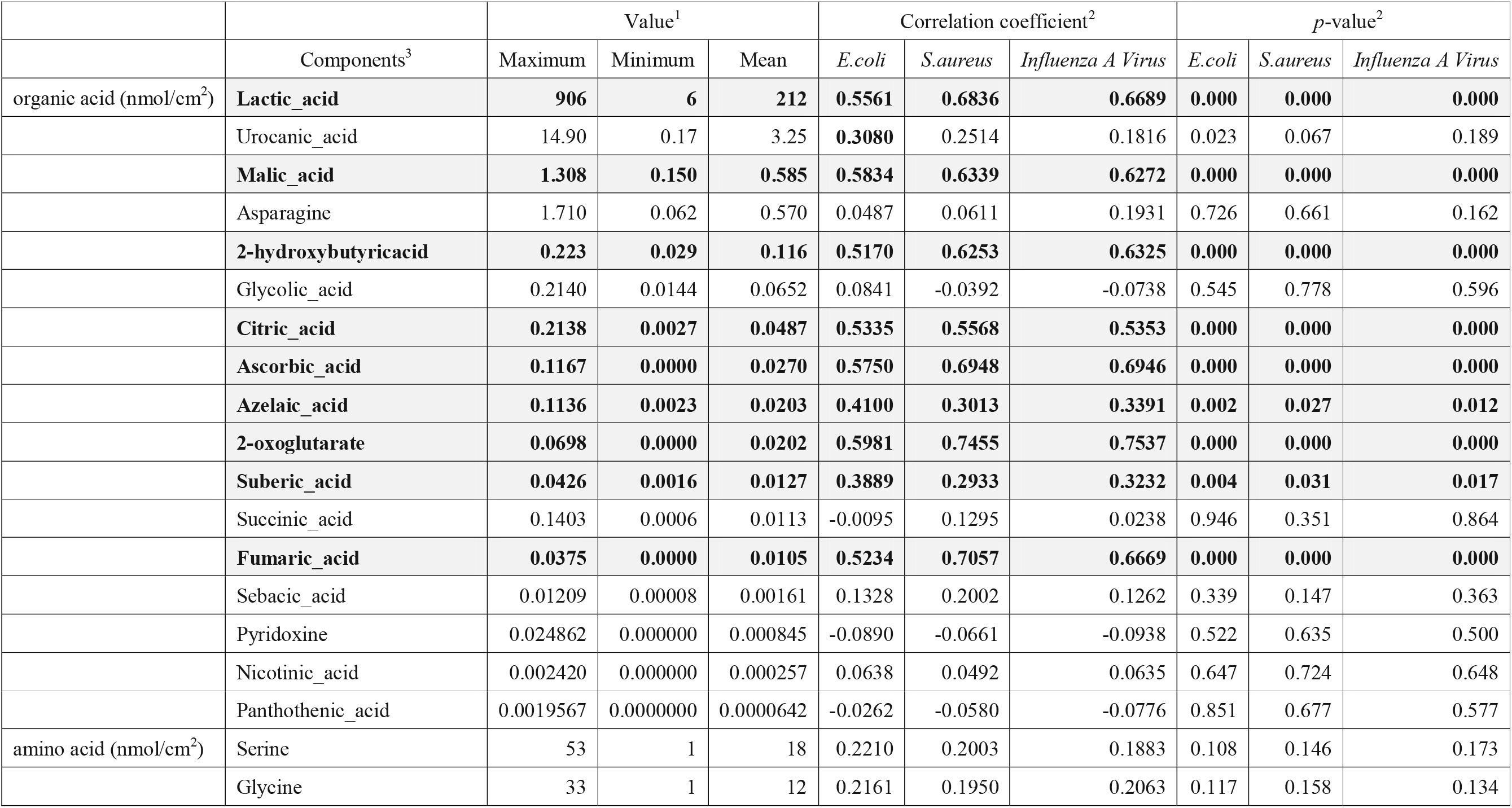
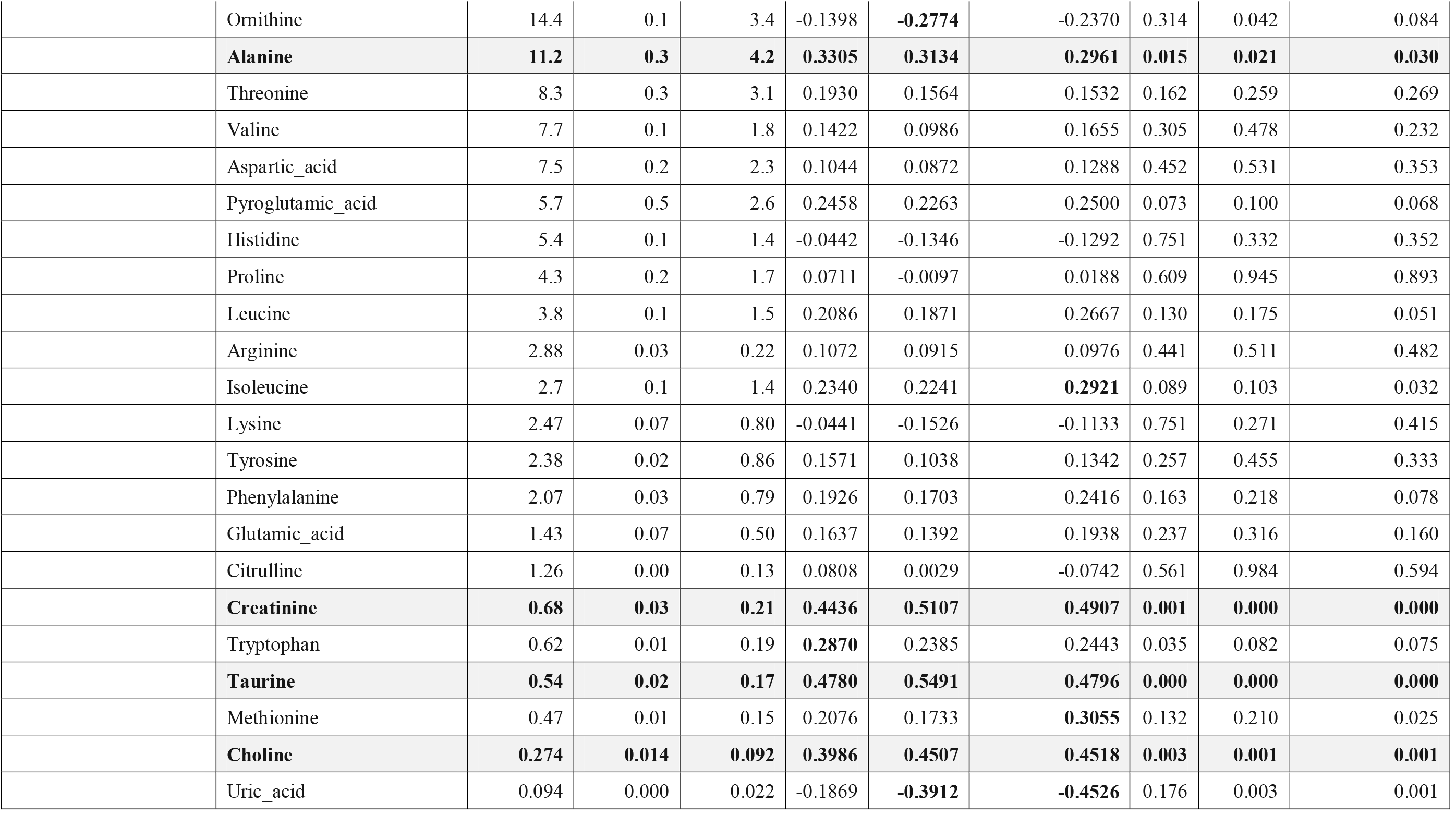
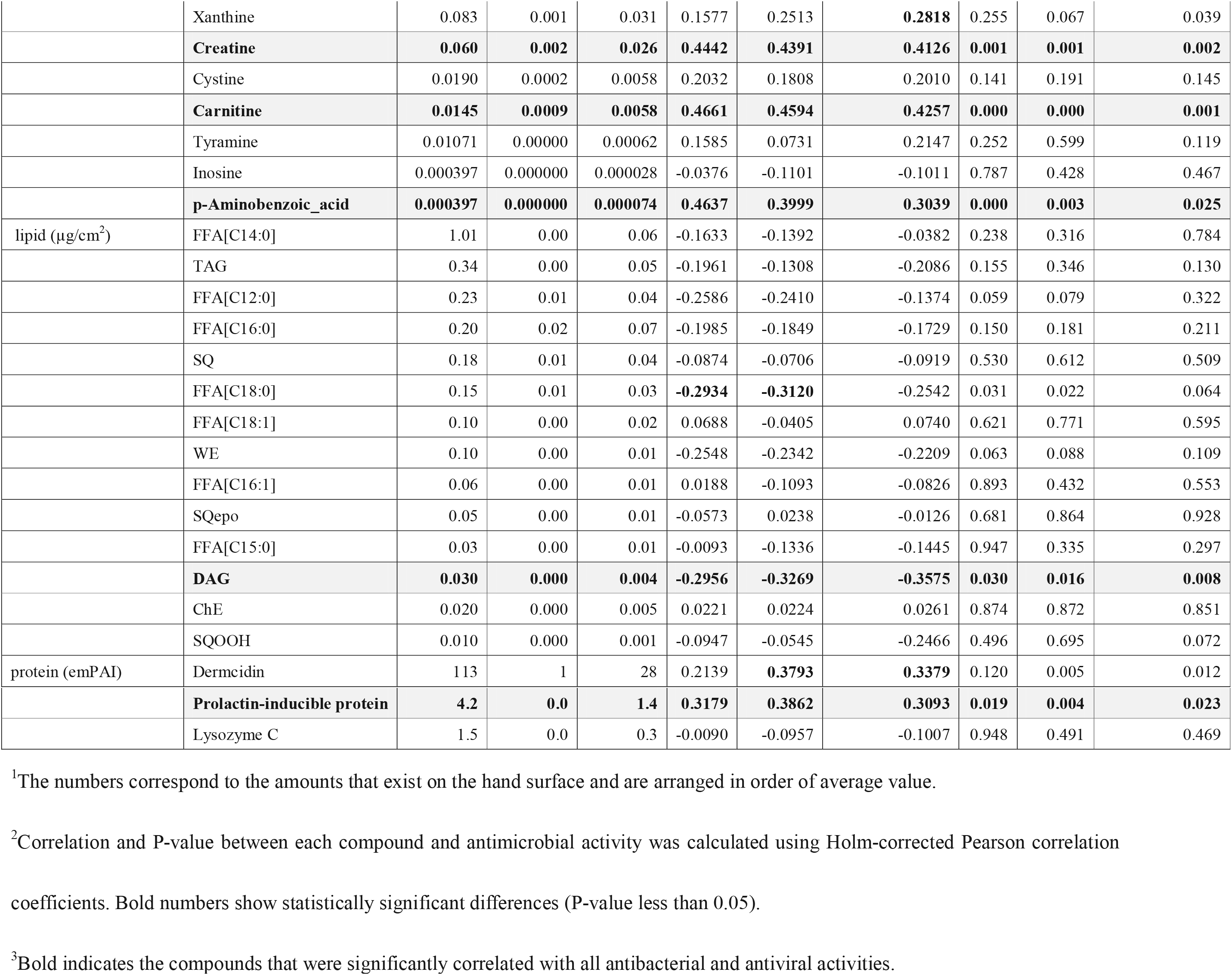
Relationship between hand surface components and antimicrobial activity.

### 3.4. Verification of the effect of lactic acid application on the antimicrobial activity of hands

Lactic acid is abundant on the hands and is highly correlated with the antimicrobial activity of the surface layer component. In fact, lactic acid has also been shown to have antimicrobial activity in test tube experiments [17]. However, these results are based on the evaluation of liquid contact, and it is unclear whether the amount of lactic acid in this study can enhance the levels of antimicrobial activity on hands. Therefore, based on the amount of lactic acid found for the 54 participants in this study (Figure 4A), a lactic acid solution was added to the hands of four subjects to verify whether it improved the antimicrobial activity. To reduce the contribution of natural lactic acid to the skin, the subjects washed their hands just before the experiment. Immediately after the hand washing, we confirmed that the amount of lactic acid on the hands was <30 nmoL/cm^2^ in each subject (Figure 4B). Although individual differences were observed in all four subjects, an improvement in antimicrobial activity was confirmed within the range of the amount of lactic acid on the hands, suggesting that lactic acid is important for hand antimicrobial activity. Furthermore, hydrochloric acid (HCl) solution with an equivalent pH (pH = 2.24) to that of the maximum amount of lactic acid in Figure 4B (652 mol/cm^2^) was applied to the hands of subjects S2 and S4. As a result, HCl was also found to improve the antimicrobial activity (*P* < 0.05), although not as much as lactic acid (*P* < 0.05) (Figure S4).

**Figure 4.**
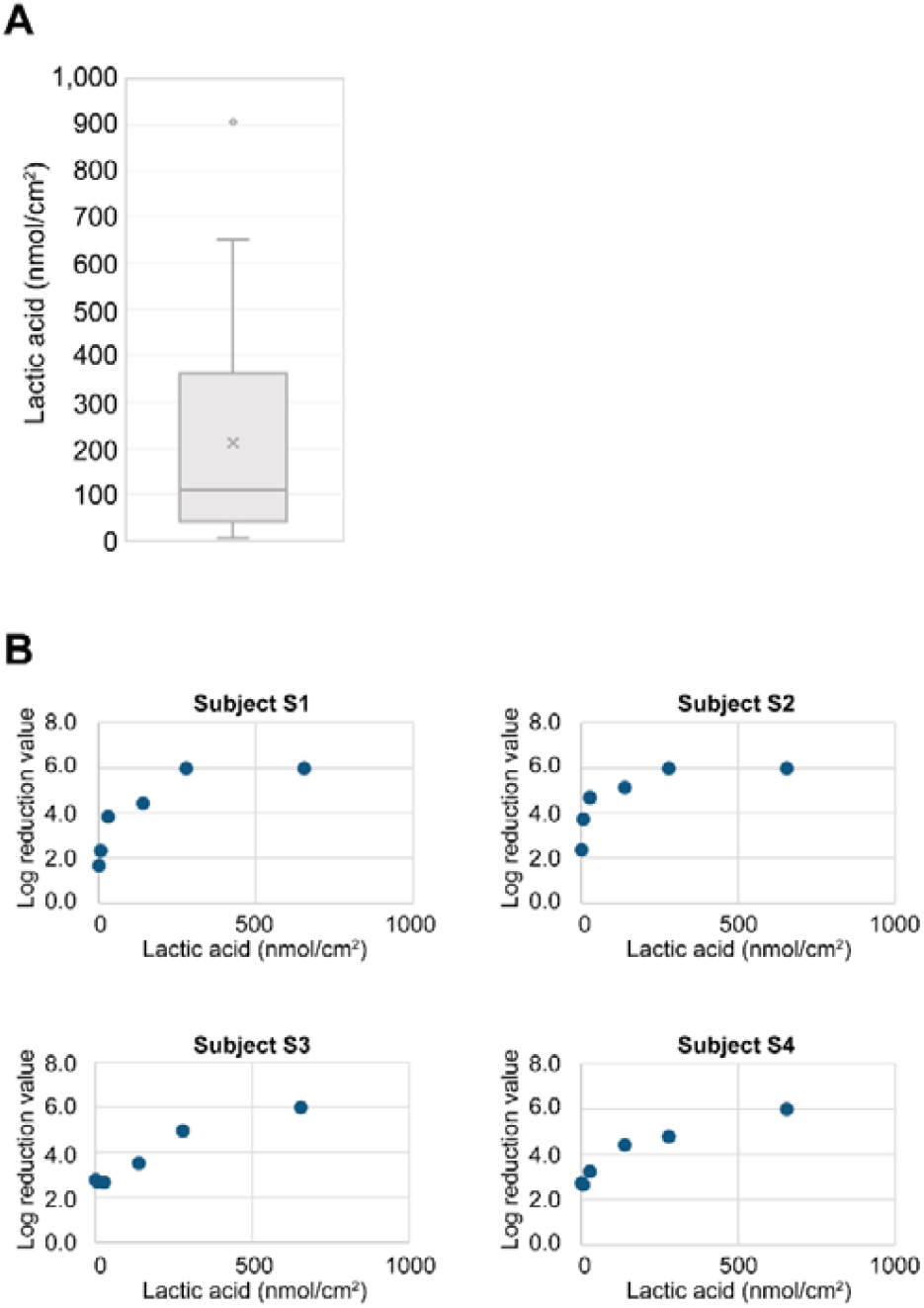
Verification of the effect of lactic acid application on the antimicrobial activity of hands. A: Amount of lactic acid on the surface of the hands of the participants (N = 54). B: Bactericidal effect of applying lactic acid onto the hands of the participants. The test site was 2 × 2 cm on the palm. After hand washing, lactic acid aqueous solution was added the test site, followed by drying for 5 min. Then, 10 µl of the bacterial solution (*E. coli*, OD = 1) was added to the test site, spread for 30 s, followed by drying for 3 min. The log reduction value denotes the relative logarithmic reduction of viable bacteria.

## 4. Discussion

In this study, we developed a novel methodology to quantify and characterize the function of the surface infection barrier on the hands that inactivates bacteria and viruses. For the first time, we demonstrated that the infection barrier found on the surface of hands plays a crucial role in quantifying the risk of the transmission of infectious diseases. In addition, we identified a list of components found on the hand surface that was correlated with antimicrobial activity, and demonstrated that lactic acid is a key component of this infection barrier.

We confirmed the reliability of these methods by identifying a positive correlation between the *in vitro* and *in vivo* methods in which bacteria are applied to hands. The data suggest that surface components are associated with the antimicrobial activity of hands. However, the correlation coefficient value was not high (r = 0.66, *P* = 0.1), suggesting that the antimicrobial activity of hands is not only due to the surface components but also to other factors, especially skin properties. such as pH and temperature [18].

Interestingly, in this study, despite exhibiting higher antimicrobial activity on the hands, the low morbidity group conducted hand washing and sanitizing less frequently than the high morbidity group. This may simply be due to a greater hygiene awareness in the high morbidity group, since these individuals are more susceptible to infection. Although there is no doubt that hand washing and sanitizing are effective for the instantaneous removal of pathogens [19], these actions cannot control the high frequency at which individuals unconsciously touch their face or potentially contaminated objects in their immediate environment. It is worth noting that the subjects in this study, who were part of the general public, appeared to have a lower hygiene awareness than medical professionals. This suggests that the inherent antimicrobial activity of hands plays a role in preventing infections *via* unconscious transmission.

In addition, our study showed that not only the antimicrobial activity on hands, but also oral mucosal moisture (and saliva volume) was associated with infection prevalence. Oral mucosal moisture is used as an objective indicator of the amount of saliva secretion [20], and a similar tendency was observed in the actual saliva volume. Saliva exerts a protective function against infection and is known to be important in preventing attack by pathogens [21, 22]. The results of this study strongly suggest that not only the hands, but also the oral cavity, exert a first-line of defense against bacterial and viral infections, and thus are important for preventing infection. On the other hand, there was no significant difference in NK cell activity and nasal mucosa clearance between the low morbidity and high morbidity groups. Many studies have shown that these biological functions are important in the prevention of infectious diseases [23, 24]. This study was conducted for subjects living with children under elementary school age, which is an environmental factor that greatly affects the prevalence of infectious diseases [25]. Limiting the attributes of the participants, the two groups in this study had similar lifestyles, such as commuting style, working style, sleep time, and stress level, among others. Thus, the population studied is unlikely to make a difference in the levels of NK cell activity and nasal mucosa clearance. The main limitation of this retrospective study is the small size of the study. Large-scale and prospective trials will be needed to better clarify the association between the antimicrobial activity of hands and morbidity.

Next, a comprehensive analysis was performed to identify the factors that control hand surface barrier activity. In previous study, sebum-containing components, especially fatty acids, on the skin were found to exhibit antimicrobial properties [29]. However, none had a significant positive correlation with the antimicrobial activity of the hands, and only a negative correlation with DAG. Compared with previous reports, the amount of lipid detected was very small, and since there are no sebaceous glands in the palm, it is highly likely that the lipids analyzed were mainly derived from sources other than the hands. On the other hand, prolactin-inducible protein was significantly correlated with antimicrobial activities among the water-soluble antimicrobial proteins, and was suggested to also be involved in the surface infection barrier of the hands. Furthermore, several amino acids were also found to be positively correlated, however, there are no prior report related to their antimicrobial properties, and the details underyling this action remain unknown.

Lactic acid is prominently present on hands. Although the hands do not have any sebaceous glands, there are many sweat glands (518/cm^2^) compared to other sites [26]. In addition to lactic acid, several organic acids were also highly correlated with antimicrobial activity, which are known to exert antimicrobial activity [27]. In general, organic acids are non-dissociative and pass through the cell membrane of the bacterium, before becoming dissociated and releasing protons to damage the bacterium from within [28]. The antimicrobial properties of organic acids depend on their pH [17]. Interestingly, almost all organic acids that exhibit a significant correlation with antimicrobial activity are key metabolites of metabolic pathways (e.g. malic acid, fumaric acid, and citric acid). This suggests that, in addition to lowering the pH of the hands, their presence on the surface of hands may give rise to a metabolic environment that is not favorable for microbial proliferation. This is supported by the fact that lactic acid exhibits a higher antimicrobial activity than a HCl solution with a similar pH.

However, the application of lactic acid solutions raises several issues: (1) skin damage, since it is an alpha hydroxylic acid and a highly acidic solution [31]; (2) variations in the pH of the skin between individuals, ranging from 4.0 to 7.0, which would affect the efficacy of lactic acid against antimicrobial activity [30]; (3) unknown long-lasting antimicrobial effects. Therefore, a more complex formulation will need to be identified to solve these issues and enhance the antimicrobial properties of lactic acid for the development of a novel hand hygiene technology to reduce contact infection risk in daily life.

## Data Availability

The datasets used and/or analysed during the current study are available from the corresponding author on reasonable request.

## Authors’ contributions

All authors have read and approved the manuscript. YN, TM, and KM wrote the manuscript. YK and AH conducted a clinical survey to analyze the relationship between high morbidity and low morbidity to infection. KN performed the investigations related to analyzing hand surface components. YN, YO, and IM designed and executed other experiments and analyzed the data. TM and KM supervised the study and reviewed the results.

## Ethical approval

The study protocol, including sample collection, was reviewed, and approved by the Ethical Committee of the Kao Corporation. (1) and (4) were approved as S311-200807, (2) was approved as T116-18032, and (3) was approved as S108-171024. Informed written consent was obtained from all research participants after the procedures had been explained with documentation. All experiments were conducted according to the principles of the Declaration of Helsinki.

## Acknowledgements

The authors would like to thank Yuuki Yanagisawa (Kao Corp.) for their useful comments and discussions throughout the research. We also thank Akira Fuji (Kao Corp.) for their valuable discussions. This study was performed in collaboration with Chihiro Tamoto (Kao Corp.), Kaori Hayashi (Kao Corp.), Yuki Sakamoto (Kao Corp.), Toshiya Morikawa (Kao Corp.), and Yukihisa Hashimoto (Kao Corp.). Editorial support, in the form of medical writing, assembling tables, and creating high-resolution images based on authors’ detailed directions, collating author comments, copyediting, fact-checking, and referencing, was provided by Editage.

## References

1. Luby, S.P., et al., Effect of handwashing on child health: a randomised controlled trial. Lancet, 2005. 366(9481): p. 225–33.

2. Kwok, Y.L., J. Gralton, and M.L. McLaws, Face touching: a frequent habit that has implications for hand hygiene. Am J Infect Control, 2015. 43(2): p. 112–4.

3. Shiraly, R., Z. Shayan, and M.L. McLaws, Face touching in the time of COVID-19 in Shiraz, Iran. Am J Infect Control, 2020.

4. Bean, B., et al., Survival of influenza viruses on environmental surfaces. J Infect Dis, 1982. 146(1): p. 47–51.

5. van Doremalen, N., et al., Aerosol and Surface Stability of SARS-CoV-2 as Compared with SARS-CoV-1. N Engl J Med, 2020. 382(16): p. 1564–1567.

6. White, C.G., et al., Reduction of illness absenteeism in elementary schools using an alcohol-free instant hand sanitizer. J Sch Nurs, 2001. 17(5): p. 258-65.

7. Aiello, A.E. and E. Larson, Antibacterial cleaning and hygiene products as an emerging risk factor for antibiotic resistance in the community. Lancet Infect Dis, 2003. 3(8): p. 501–6.

8. Lingaas, E. and M. Fagernes, Development of a method to measure bacterial transfer from hands. J Hosp Infect, 2009. 72(1): p. 43–9.

9. Schauber, J. and R.L. Gallo, Antimicrobial peptides and the skin immune defense system. J Allergy Clin Immunol, 2009. 124(3 Suppl 2): p. R13–8.

10. Turner, R.B., et al., Efficacy of organic acids in hand cleansers for prevention of rhinovirus infections. Antimicrob Agents Chemother, 2004. 48(7): p. 2595–8.

11. Iketani, R., et al., The Effect of Tea Catechins on Natural Killer Cell Activity in the Elderly: A Pilot Study. Clinical Pharmacology, 2019. 50(4): p. 139–145.

12. Ito, J.T., et al., Nasal Mucociliary Clearance in Subjects With COPD After Smoking Cessation. Respir Care, 2015. 60(3): p. 399–405.

13. Ishii, S., et al., Identification of the Catechin Uptake Transporter Responsible for Intestinal Absorption of Epigallocatechin Gallate in Mice. Sci Rep, 2019. 9(1): p. 11014.

14. Ishihama, Y., et al., Exponentially modified protein abundance index (emPAI) for estimation of absolute protein amount in proteomics by the number of sequenced peptides per protein. Mol Cell Proteomics, 2005. 4(9): p. 1265–72.

15. Han, X., K. Yang, and R.W. Gross, Multi-dimensional mass spectrometry-based shotgun lipidomics and novel strategies for lipidomic analyses. Mass Spectrom Rev, 2012. 31(1): p. 134–78.

16. Fukui, S., et al., A mild hand cleanser, alkyl ether sulphate supplemented with alkyl ether carboxylic acid and alkyl glucoside, improves eczema on the hand and prevents the growth of Staphylococcus aureus on the skin surface. Int J Cosmet Sci, 2016. 38(6): p. 599–606.

17. Kim, S.A. and M.S. Rhee, Marked synergistic bactericidal effects and mode of action of medium-chain fatty acids in combination with organic acids against Escherichia coli O157:H7. Appl Environ Microbiol, 2013. 79(21): p. 6552–60.

18. Maliyar, K., R. Persaud-Jaimangal, and R.G. Sibbald, Associations Among Skin Surface pH, Temperature, and Bacterial Burden in Wounds. Adv Skin Wound Care, 2020. 33(4): p. 180–185.

19. Aiello, A.E., et al., Effect of hand hygiene on infectious disease risk in the community setting: a meta-analysis. Am J Public Health, 2008. 98(8): p. 1372–81.

20. Takahashi, F., T. Koji, and O. Morita, The Usefulness of an Oral Moisture Checking Device (Moisture Checker for Mucus^R^). Nihon Hotetsu Shika Gakkai Zasshi, 2005. 49(2): p. 283–289.

21. Heo, S.M., et al., Host defense proteins derived from human saliva bind to Staphylococcus aureus. Infect Immun, 2013. 81(4): p. 1364–73.

22. Limsuwat, N., et al., Sialic acid content in human saliva and anti-influenza activity against human and avian influenza viruses. Arch Virol, 2016. 161(3): p. 649–56.

23. Wilson, R., et al., Upper respiratory tract viral infection and mucociliary clearance. Eur J Respir Dis, 1987. 70(5): p. 272–9.

24. Hazeldine, J. and J.M. Lord, The impact of ageing on natural killer cell function and potential consequences for health in older adults. Ageing Res Rev, 2013. 12(4): p. 1069–78.

25. Sakurai, R., et al., Long-term effects of an intergenerational program on functional capacity in older adults: Results from a seven-year follow-up of the REPRINTS study. Arch Gerontol Geriatr, 2016. 64: p. 13–20.

26. Taylor, N.A. and C.A. Machado-Moreira, Regional variations in transepidermal water loss, eccrine sweat gland density, sweat secretion rates and electrolyte composition in resting and exercising humans. Extrem Physiol Med, 2013. 2(1): p. 4.

27. Brul, S. and P. Coote, Preservative agents in foods. Mode of action and microbial resistance mechanisms. Int J Food Microbiol, 1999. 50(1-2): p. 1–17.

28. Mani-López, E., H.S. García, and A. López-Malo, Organic acids as antimicrobials to control Salmonella in meat and poultry products. Food Research International, 2012. 45(2): p. 713–721.

29. Takigawa, H., et al., Deficient production of hexadecenoic acid in the skin is associated in part with the vulnerability of atopic dermatitis patients to colonization by Staphylococcus aureus. Dermatology, 2005. 211(3): p. 240–8.

30. Lambers, H., et al., Natural skin surface pH is on average below 5, which is beneficial for its resident flora. Int J Cosmet Sci, 2006. 28(5): p. 359–70.

31. Tang, S.C. and J.H. Yang, Dual Effects of Alpha-Hydroxy Acids on the Skin. Molecules, 2018. 23(4).

