## Supplemental file for "Detailed Analysis of Surface Infection Barrier on Hands: Relationship with Morbidity to Infection Diseases and Identification of Antimicrobial Components"

**Supplementary files**

**Table S1. Participant characteristics**

| **Characteristics** | **High morbidity group** | **Low morbidity group** |
| --- | --- | --- |
| n | 55 | 54 |
| Age (years), mean (SD) | 39.9 (4.9) | 40.6 (5.4) |
| Sex, n (%) |  |  |
| Men | 17 | 21 |
| Women | 38 | 33 |
| Number of flu within 3 years | 2 times or more | 0 times |
| Number of colds within a year (Cough, runny nose, sore throat, etc. with fever of 37.5 ℃ or higher) | 3 times or more | 0 times |

**Table S2. Questionnaire regarding lifestyles and hygiene behaviors**

| Questionnaire | Answer | Low morbidity group (N=54) | | High morbidity group (N=55) | | *P*-value^1^ |
| --- | --- | --- | --- | --- | --- | --- |
|  |  | Numbers^1^ | Percentage^1^ | Numbers^1^ | Percentage^1^ |  |
| A. Regarding Life-Styles | | | | | |  |
| 1) Are you commuting by train or bus? | Yes | 22 | 41% | 29 | 53% | 0.210 |
|  | No | 32 | 59% | 26 | 47% | 0.210 |
| 2) Do you often use public facilities (stations, libraries, movie theaters, sports facilities, parks, etc.) (twice or more / week)? | Yes | 29 | 54% | 29 | 53% | 0.919 |
|  | No | 25 | 46% | 26 | 47% | 0.919 |
| 3) Do you have a lot of desk work (5 hours or more / day) | Yes | 22 | 41% | 17 | 31% | 0.284 |
|  | No | 32 | 59% | 38 | 69% | 0.284 |
| 4) Do you smoke? | Smoking | 7 | 13% | 6 | 11% | 0.741 |
|  | I smoked in the past | 12 | 22% | 18 | 33% | 0.741 |
|  | I have never smoked | 35 | 65% | 31 | 56% | 0.367 |
| 5) How often do you drink alcohol? | Almost every day | 9 | 17% | 4 | 7% | 0.130 |
|  | within 5 days / week | 9 | 17% | 11 | 20% | 0.653 |
|  | within 2 days / week | 14 | 26% | 16 | 29% | 0.711 |
|  | Almost never drink | 22 | 41% | 24 | 44% | 0.760 |
| 6) How often do you exercise for 30 minutes or more at a time (including commuting such as walking or biking) | Almost every day | 6 | 11% | 14 | 25% | 0.053 |
|  | within 5 days / week | 2 | 4% | **10** | **18%** | **0.016** |
|  | within 2 days / week | 19 | 35% | 14 | 25% | 0.269 |
|  | Almost never exercise | **27** | **50%** | 17 | 31% | **0.042** |
| 7) Do you have enough sleep | Yes | 40 | 74% | 37 | 67% | 0.436 |
|  | No | 14 | 26% | 18 | 33% | 0.436 |
| 8) Do you try to have a nutritionally balanced diet? | Yes | 42 | 78% | 47 | 85% | 0.301 |
|  | No | 12 | 22% | 8 | 15% | 0.301 |
| 9) Are you able to release stress? | Yes | 37 | 69% | 32 | 58% | 0.263 |
|  | No | 17 | 31% | 23 | 42% | 0.263 |
| B. Regarding Hygiene Behaviors | | | | | |  |
| 10) How often do you wash your hands? | Almost every day | 44 | 81% | **53** | **96%** | 0.013 |
|  | Sometimes (only during infection season) | 7 | 13% | 2 | 4% | 0.077 |
|  | Nothing | 3 | 6% | 0 | 0% | 0.077 |
| 11) How often do you use hand sanitizer? | Almost every day | 1 | 2% | **9** | **16%** | **0.009** |
|  | Sometimes (only during infection season) | 24 | 44% | 34 | 62% | 0.069 |
|  | Nothing | **29** | **54%** | 12 | 22% | **0.001** |
| 12) How often do you gargle? | Almost every day | 25 | 46% | **41** | **75%** | **0.003** |
|  | Sometimes (only during infection season) | **24** | **44%** | 12 | 22% | 0.012 |
|  | Nothing | 5 | 9% | 2 | 4% | 0.231 |
| 13) How often do you wear mask? | Almost every day | 2 | 4% | 7 | 13% | 0.087 |
|  | Sometimes (only during infection season) | 35 | 65% | **46** | **84%** | **0.025** |
|  | Nothing | **17** | **31%** | 2 | 4% | **0.000** |
| 14) How much do you get the flu vaccine? | 2 times every year | 0 | 0% | 2 | 4% | 0.157 |
|  | 1 time every year | 0 | 0% | **32** | **59%** | **0.000** |
|  | Once every two years | 0 | 0% | **8** | **15%** | **0.000** |
|  | I have not been vaccinated in the last few years | **29** | **54%** | 5 | 9% | **0.000** |
|  | I have never / do not remember being vaccinated | **25** | **46%** | 8 | 15% | **0.000** |

^1^The significance test was performed with the null hypothesis. Bold numbers show statistically significant high (the p-value less than 0.05).

**Table S3. Results calculated by the decision tree analysis**

| Parameters | reference value calculated by the decision tree analysis | *P*-value | OR | 95% CI |
| --- | --- | --- | --- | --- |
| Antimicrobial activity of hand for *E. coli* | 0.154 | < 0.001 | 7.2 | 2.64 - 19.50 |
| oral mucosal moisture | 27.7 | 0.004 | 5.7 | 2.08 - 15.60 |
| age | 41 | 0.033 | 0.12 | 0.01 - 1.08 |
|  | 38 | 0.209 | 2.86 | 0.53 - 15.41 |

**Figure S1. Sites for the collection of surface components for compounds analysis (left hand) and antimicrobial analysis (right hand).**

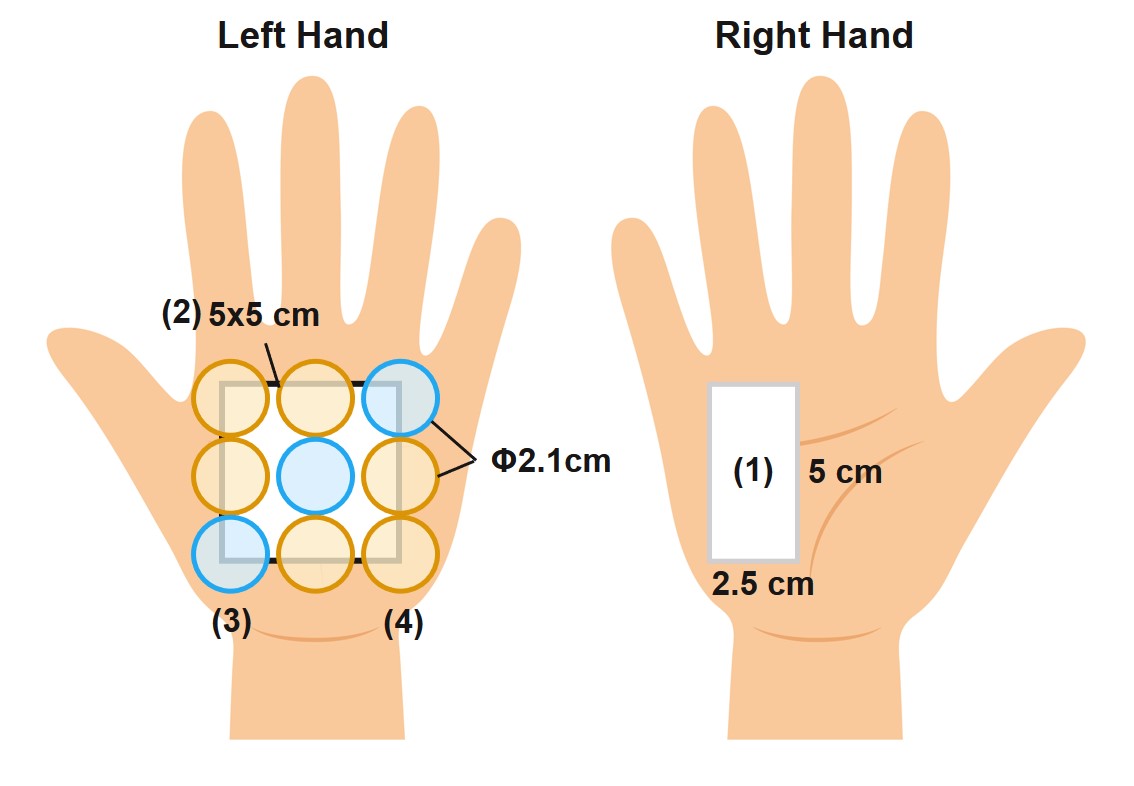

For the collection of water-soluble components, 9 sheets of filter paper were attached to the palm of the left hand for 5 minutes, and 60 µL of ultrapure water was added to each filter paper. The filter paper was divided into 3 sheets (for low molecular weight compounds) and 6 sheets (for antibacterial peptides). For lipid collection on the skin surface, the left palm was scraped with a swab soaked in ethanol and collected in a screw tube (2). For antimicrobial analysis, the palm of right hand was rubbed with a swab soaked in 50% ethanol. The tip of a swab was cut off and 600 µL of 50% ethanol were added (1).

**Figure S2. Results immediately after washing the hands of a person with high antimicrobial activity (Figure 1A left).**

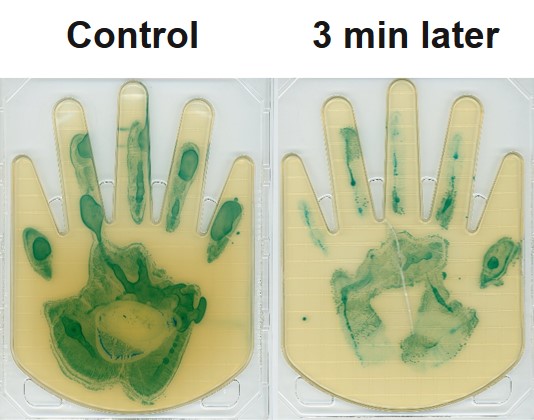

The results show 30 seconds as the control and 3 minutes after applying *E.coli* solution of OD= 0.2 to the hand.

**Figure S3. Relationship between the amount of lactic acid on hand surface and the antimicrobial activities of hand surface components.**

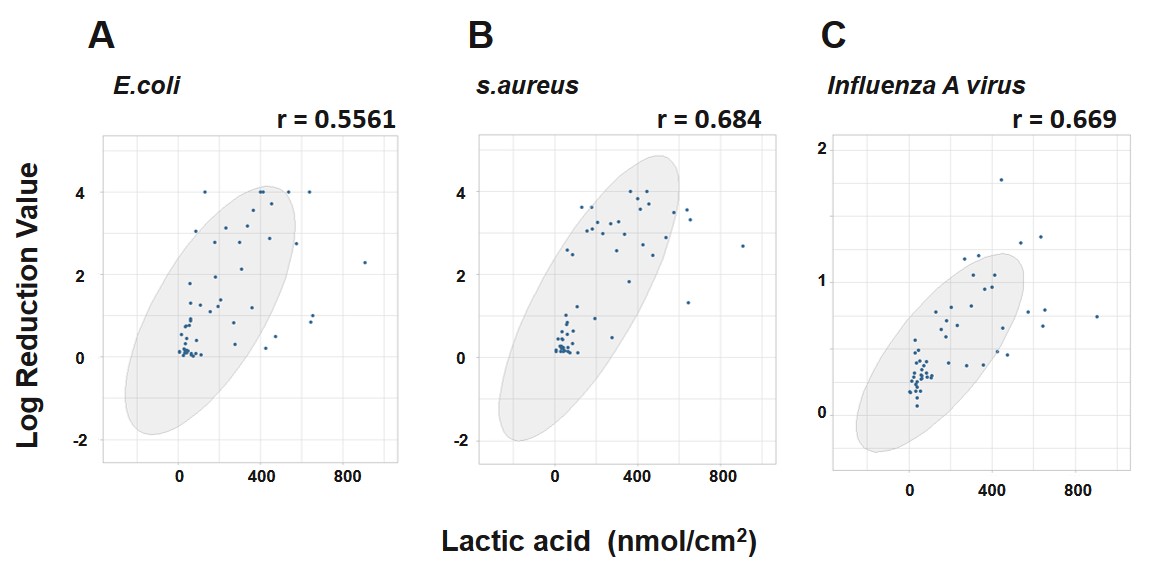

The horizontal axis shows the amount of lactic acid on hands, and the vertical axis shows the antimicrobial activities. Antimicrobial activities were measured using *in vitro* method and log reduction value means the relative logarithmic reduction of bacterial or viral number. A, B and C show the results against *E.coli*, *S.aureus* and Influenza A virus.

**Figure S4. Comparison of bactericidal activities on the hand by application of hydrochloric acid solution with the same pH as lactic acid.**

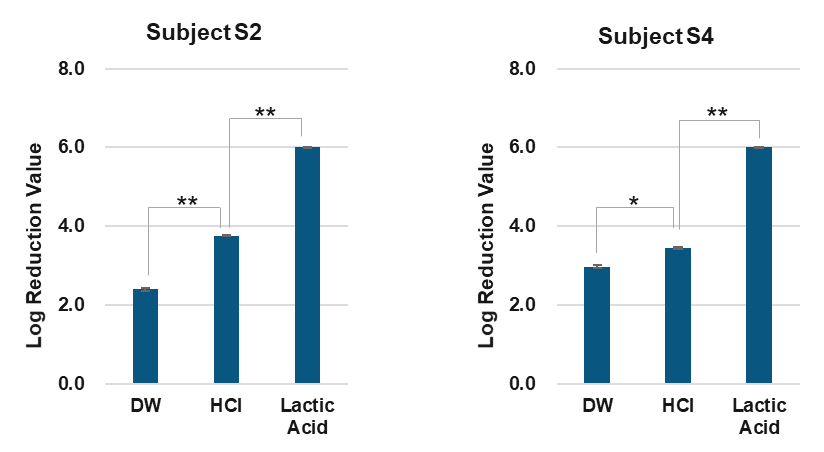

Lactic acid aqueous solution or hydrochloric acid solution (HCl) was added to the test site, spread over, and dried. Next, bacterial solution (*E.coli*, OD=1) was added to the test site, spread, and dried. Log reduction value means the relative logarithmic reduction of viable bacteria. Hydrochloric acid solution (HCl) was adjusted to 2.24 with the same pH as lactic acid. Asterisks indicate significant difference between two samples (t test, **P<0.01　and *P<0.05
